# Combined Assessment of Pulmonary Artery Pulsatility Index and Compliance for Risk Stratification in Advanced Heart Failure: A Post-hoc Analysis of the ESCAPE Trial

**DOI:** 10.1101/2025.08.25.25334407

**Authors:** Yuta Ozaki, Yusuke Uemura, Toru Kondo, Shingo Kazama, Shogo Yamaguchi, Takashi Okajima, Takayuki Mitsuda, Shinji Ishikawa, Kenji Takemoto, Takahiro Okumura, Masato Watarai, Toyoaki Murohara

**Affiliations:** Cardiovascular Center, Anjo Kosei Hospital, Anjo, Japan; Department of Cardiology, Nagoya University Graduate School of Medicine, Nagoya, Japan

**Keywords:** advanced heart failure, pulmonary artery pulsatility index, pulmonary arterial capacitance, prognosis, risk stratification

## Abstract

**Background:** In advanced heart failure (HF), dynamic interplays between right ventricular (RV) dysfunction and afterload complicate prognostic assessments. The pulmonary artery pulsatility index (PAPi) reflects RV function, while pulmonary arterial capacitance (PAC) represents the pulsatile component of afterload. We investigated whether a combined PAPi-PAC assessment improves risk stratification in patients with decompensated advanced HF.

**Methods:** This retrospective analysis included 187 patients with advanced HF and complete baseline PAPi and PAC data from the ESCAPE trial. The composite endpoint included all-cause death, left ventricular assist device implantation, or orthotopic heart transplantation within 6 months. A grid search approach identified optimal cutoffs for PAPi (2.00) and PAC (1.64), stratifying patients into four groups.

**Results:** In Cox models adjusted for the Get With The Guidelines-Heart Failure (GWTG-HF) score, the low PAPi/low PAC group had the highest risk of the composite endpoint compared with the high PAPi/high PAC group (hazard ratio [HR] 3.167, 95% confidence interval [CI] 1.387–7.230), followed by the high PAPi/low PAC (HR 2.388, 95%CI 1.075–5.305) and the low PAPi/high PAC groups (HR 2.266, 95%CI 0.962–5.334). Model discrimination—assessed using C-indices—was improved by the addition of both PAPi and PAC to the GWTG-HF score (C-index 0.688 vs. 0.652; ΔC-index +0.047, 95%CI 0.002– 0.100, P=0.038). Conversely, models incorporating PAPi or PAC alone did not significantly enhance discrimination.

**Conclusions:** In patients with decompensated advanced HF, early invasive hemodynamic phenotyping using combined PAPi-PAC enhances risk stratification beyond established clinical scores and may aid timely consideration of advanced therapies in high-risk individuals.

**Clinical Perspective:** *What is New?:*

- Models incorporating both the pulmonary artery pulsatility index (PAPi) and pulmonary arterial capacitance (PAC) to the Get With The Guidelines-Heart Failure score demonstrated enhanced discriminatory ability.
- Incorporation of only PAPi or PAC alone did not significantly improve model discrimination.

*What Are the Clinical Implications?:*

- In patients with decompensated advanced HF, early invasive hemodynamic phenotyping using a combined PAPi-PAC approach enhances risk stratification.
- Combined PAPi-PAC assessment may aid decision-making regarding advanced therapies in high-risk individuals.

## Introduction

Despite advances in heart failure (HF) management, a significant number of patients progress to advanced disease (1, 2). The prevalence of advanced HF is increasing, driven by an aging population, improved survival in earlier stages, and the broader adoption of guideline-directed medical therapy (2). Nevertheless, the prognosis remains poor, with a 1-year mortality rate of approximately 50% (3).

Right ventricular (RV) dysfunction—a prognostic indicator and key component of disease progression in advanced HF (4)—frequently develops owing to sustained elevation of left-sided filling pressure, which induces pulmonary vascular remodeling and leads to reduced pulmonary arterial capacitance (PAC) (5–7). Given this pathophysiological interplay, assessment of RV function should be interpreted in the context of PAC for a more comprehensive evaluation of RV performance and afterload.

The pulmonary artery pulsatility index (PAPi), calculated as the pulmonary artery pulse pressure divided by the right atrial pressure, is an invasively assessed hemodynamic parameter that reflects RV function (8). While it is widely used to predict RV failure and adverse outcomes in patients who have undergone left ventricular assist device implantation, recent studies have highlighted its broader prognostic utility in various HF populations, including those with advanced disease (9, 10). PAC, defined as the stroke volume divided by the pulmonary artery pulse pressure, represents the pulsatile component of RV afterload and is particularly sensitive to elevated left-sided filling pressure (11). We have previously reported that the combined assessment of PAPi and PAC offers an effective framework for prognostic stratification and hemodynamic phenotyping in patients with compensated HF, both with preserved and reduced ejection fractions (12, 13). However, whether this combined hemodynamic classification also holds prognostic value in advanced HF remains unclear.

In the present study, we analyzed data from the North American Evaluation Study of Congestive Heart Failure and Pulmonary Artery Catheterization Effectiveness (ESCAPE) trial to investigate whether baseline PAPi and PAC are associated with clinical outcomes in patients hospitalized for decompensated advanced HF (14). Since these parameters were collected during early hospitalization and reflect the acute hemodynamic state, this analysis aimed to clarify their prognostic relevance at the time of the initial invasive assessment, independent of in-hospital treatment response or discharge condition.

## Methods

### Study Population

Herein, we conducted a post-hoc analysis using de-identified individual-level data from the ESCAPE trial, obtained through the Biologic Specimen and Data Repository Information Coordinating Center repository of the United States National Heart, Lung, and Blood Institute. A data use agreement was signed prior to data access, and this study was exempted from institutional review board approval by the Anjo Kosei Hospital Ethics Committee (Waiver No. R24-031).

The ESCAPE trial enrolled 433 patients with advanced HF who were hospitalized across 26 centers in the United States and Canada between January 2000 and November 2003. The trial design and main results have been described previously (14). Briefly, patients with severe symptomatic HF despite recommended therapy were randomized to receive either clinical assessment alone or pulmonary artery catheterization-guided therapy. Enrolled patients met the following inclusion criteria: a left ventricular ejection fraction ≤30%, persistent symptoms despite guideline-directed medical therapy for at least 3 months, a systolic blood pressure ≤125 mmHg, and at least one clinical sign or symptom of congestion. In addition, patients were required to meet at least one of the following indicators of advanced HF severity: a HF-related hospitalization within the previous year, an urgent emergency department visit, or use of high-dose loop diuretics (furosemide >160 mg/day or equivalent) within the prior month. Major exclusion criteria included a serum creatinine level >3.5 mg/dL, recent or ongoing use of high-dose inotropes, or any use of milrinone during the index hospitalization.

Patients in the pulmonary artery catheterization-guided arm were managed with the aim of achieving target hemodynamics of pulmonary capillary wedge pressure ≤15 mmHg or right atrial pressure ≤8 mmHg, in addition to symptom-based goals used in both groups. Use of intravenous inotropes for routine management was discouraged during hospitalization.

For this post-hoc analysis, we included patients for whom invasive hemodynamic data required to calculate both baseline PAPi and PAC were available, regardless of randomization arm. PAPi was calculated as the pulmonary artery pulse pressure (i.e., systolic minus diastolic pulmonary artery pressure) divided by the right atrial pressure, while PAC was defined as the stroke volume divided by the pulmonary artery pulse pressure. Because stroke volume was not directly recorded in the ESCAPE dataset, it was estimated as the cardiac output divided by the heart rate. In the ESCAPE trial, pulmonary artery catheters were placed at the time of randomization, and baseline hemodynamic parameters were collected immediately thereafter as part of the study protocol. Herein, these post-randomization baseline measurements were used for all analyses.

The endpoint was a composite of all-cause death, orthotopic heart transplantation, or left ventricular assist device implantation within 6 months of randomization.

### Statistical analysis

Normality of data distribution was assessed using the Shapiro–Wilk test. Regarding continuous variables, normally distributed data are expressed as means ± standard deviations (SDs), while non-normally distributed data are presented as medians (interquartile ranges [IQRs]). Categorical variables are summarized as counts and percentages. Group comparisons were conducted using one-way analysis of variance or Kruskal–Wallis test for continuous variables, and the chi-square test for categorical variables.

We utilized a grid search approach to determine optimal cutoffs for PAPi and PAC for predicting the composite endpoint. Each variable was dichotomized at 10-percentile intervals between the 30th and 70th percentiles, and all cutoff combinations were evaluated using Cox proportional hazards models. The concordance index (C-index) was calculated for each model, and the cutoff combination most frequently associated with the highest C-index across 10,000 bootstrap samples was selected as the final threshold.

Using the derived thresholds (PAPi, 2.00; PAC, 1.64), participants were classified into four hemodynamic groups. Kaplan–Meier survival curves were generated, which were compared using log-rank tests. Cox proportional hazards models were used to estimate hazard ratios (HRs) and 95% confidence intervals (CIs) for the composite endpoint. Multivariable models were adjusted using the Get With The Guidelines–Heart Failure (GWTG-HF) risk score, a validated clinical prediction tool originally developed to predict in-hospital mortality in patients with acute decompensated HF and subsequently validated for the prediction of long-term cardiovascular outcomes (15, 16). The score integrates variables such as age, systolic blood pressure, sodium, blood urea nitrogen, heart rate, race, and chronic obstructive pulmonary disease (15, 16).

To evaluate incremental prognostic value, the C-index of a model including only the GWTG-HF score was compared with models that additionally incorporated the dichotomized values of PAPi, PAC, or both. Changes in C-indices (ΔC-index) were estimated for each comparison using 1,000 bootstrap replicates to derive 95% CIs and P-values.

For the calculation of the GWTG-HF score, missing values in score components were imputed using multiple imputation by chained equations. Baseline characteristics were described after excluding missing values.

All statistical analyses were performed using R version 4.3.2 (R Foundation for Statistical Computing, Vienna, Austria). The following R packages were used: “*survival*”, “*survminer*”, “*purr*”, “*compareC,”* and *“mice.”* Two-sided P-values <0.05 were considered statistically significant.

## Results

Of the 433 patients enrolled in the ESCAPE trial, invasive hemodynamic data were available for 202 patients, including 187 in the pulmonary artery catheterization-guided therapy group and 15 in the standard care group who underwent right heart catheterization. Among them, complete baseline data required to calculate both the PAPi and PAC were available for 187 patients, who were subsequently included in the final analysis cohort.

In the grid search procedure to identify optimal cutoff values for PAPi and PAC, the combination of PAPi = 2.00 and PAC = 1.64 was the most frequently observed optimal threshold, being identified in 151 runs with a corresponding mean C-index of 0.710, which was therefore adopted for subsequent analyses (Figure 1).

**Figure 1.**
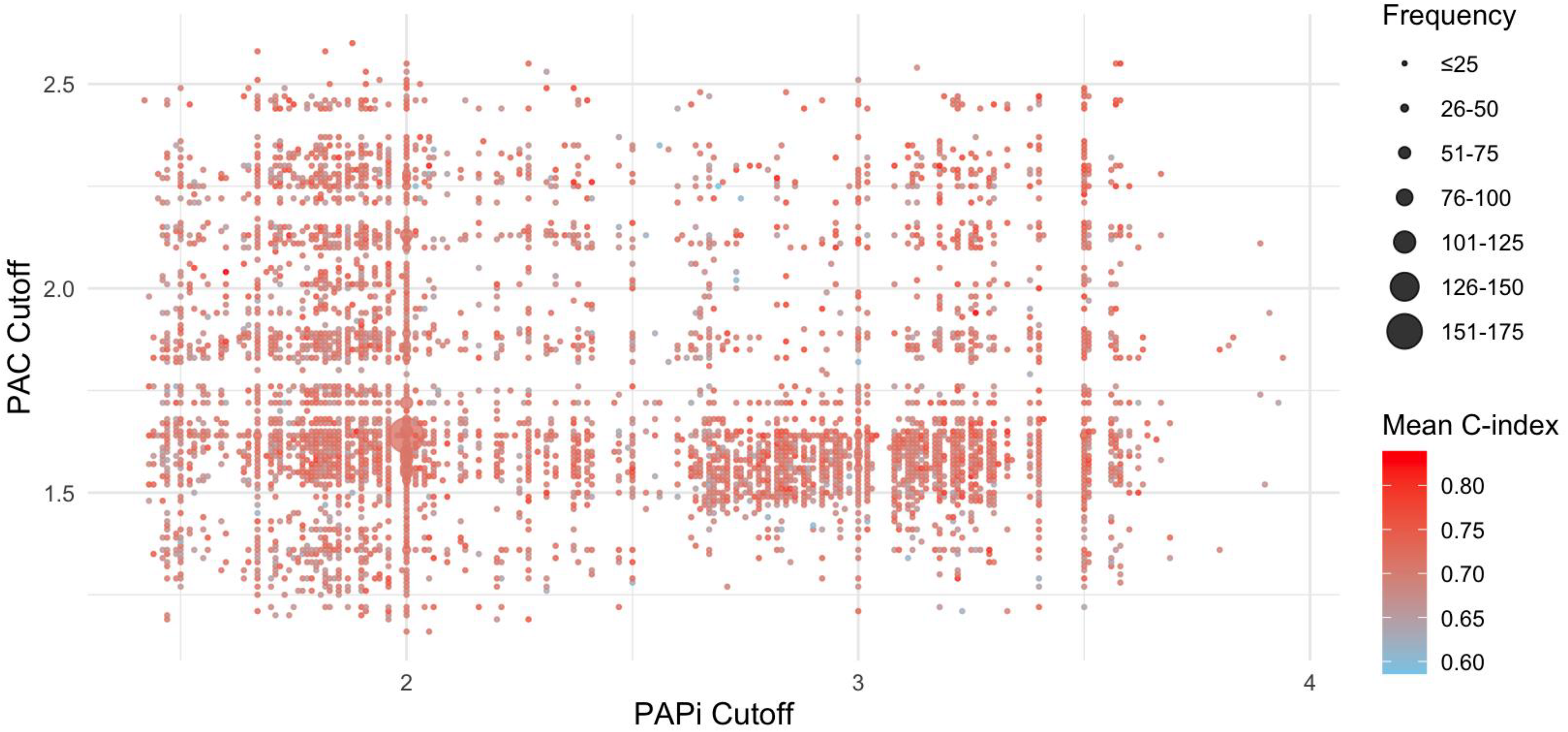
Grid search with bootstrap resampling to identify optimal cutoffs for PAPi and PAC. A bootstrap-based grid search was conducted to determine the optimal cutoff values of the PAPi and PAC for prognostic stratification. Each variable was dichotomized at 10-percentile intervals between the 30th and 70th percentiles, and all combinations were evaluated using Cox proportional hazards models across 10,000 bootstrap iterations. PAC, pulmonary arterial capacitance; PAPi, pulmonary artery pulsatility index.

Baseline clinical characteristics of the four groups defined by the PAPi-PAC classification are shown in Table 1. In the overall cohort, 49 patients (26%) were women, the mean age was 56.2 years, the median body mass index was 27.8 kg/m^2^, and the median left ventricular ejection fraction was 20.0%; these variables were comparable across the four groups. Ischemic etiology appeared to be more prevalent in the low PAPi/low PAC group compared with other groups, while the distributions of other comorbidities were largely balanced. Additionally, the low PAPi/low PAC group tended to have lower serum sodium levels and higher blood urea nitrogen concentrations. The use of both oral and intravenous HF medications was generally similar across the groups. Notably, the GWTG-HF risk score was higher in the low PAPi/low PAC group than in other groups. The median values of the PAPi and PAC were 2.33 (IQR, 1.47–3.56) and 1.72 (IQR, 1.26–2.45), respectively. Patients with both low PAPi and low PAC were characterized by higher right atrial pressure, pulmonary capillary wedge pressure, and mean pulmonary artery pressure, and lower cardiac output, whereas the low PAPi/high PAC and high PAPi/low PAC groups generally exhibited intermediate values for these parameters compared with the high PAPi/high PAC group.

**Table 1.**
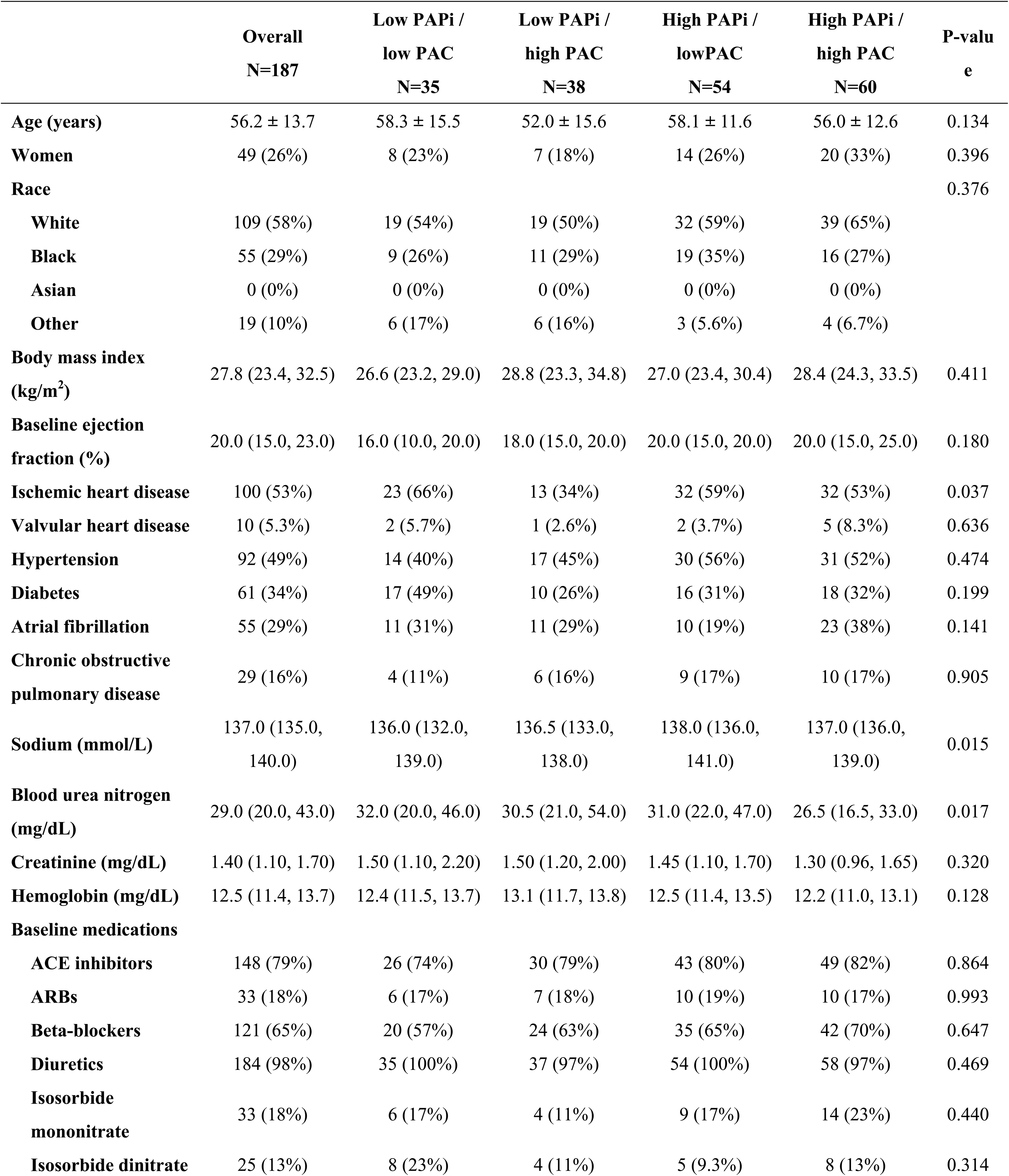
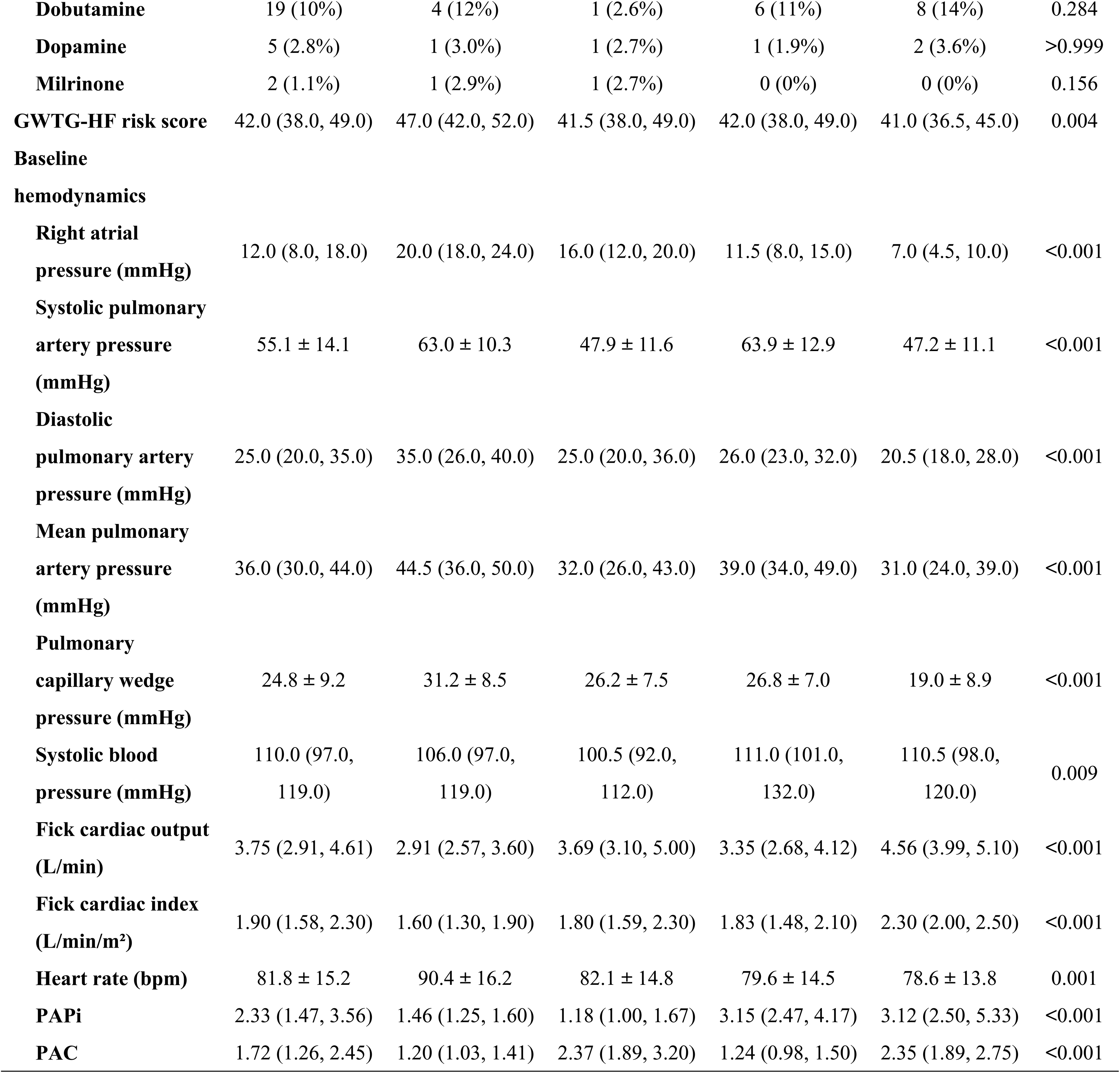
Baseline patient characteristics. Data are presented as medians (interquartile ranges), means ± standard deviations, or n (%), as appropriate. Patients were stratified into four groups based on optimally derived cutoffs for the PAPi (2.00) and PAC (1.64). P-values were calculated using one-way analysis of variance or the Kruskal–Wallis test for continuous variables, and the chi-square test for categorical variables. GWTG-HF scores were calculated after imputing missing values in its components using multiple imputation by chained equations. All other baseline characteristics in this table are described after excluding missing values. ACE, angiotensin-converting enzyme; ARBs, angiotensin receptor blockers; GWTG-HF, Get With The Guidelines-Heart Failure; PAC, pulmonary arterial capacitance; PAPi, pulmonary artery pulsatility index.

**Table 2.**
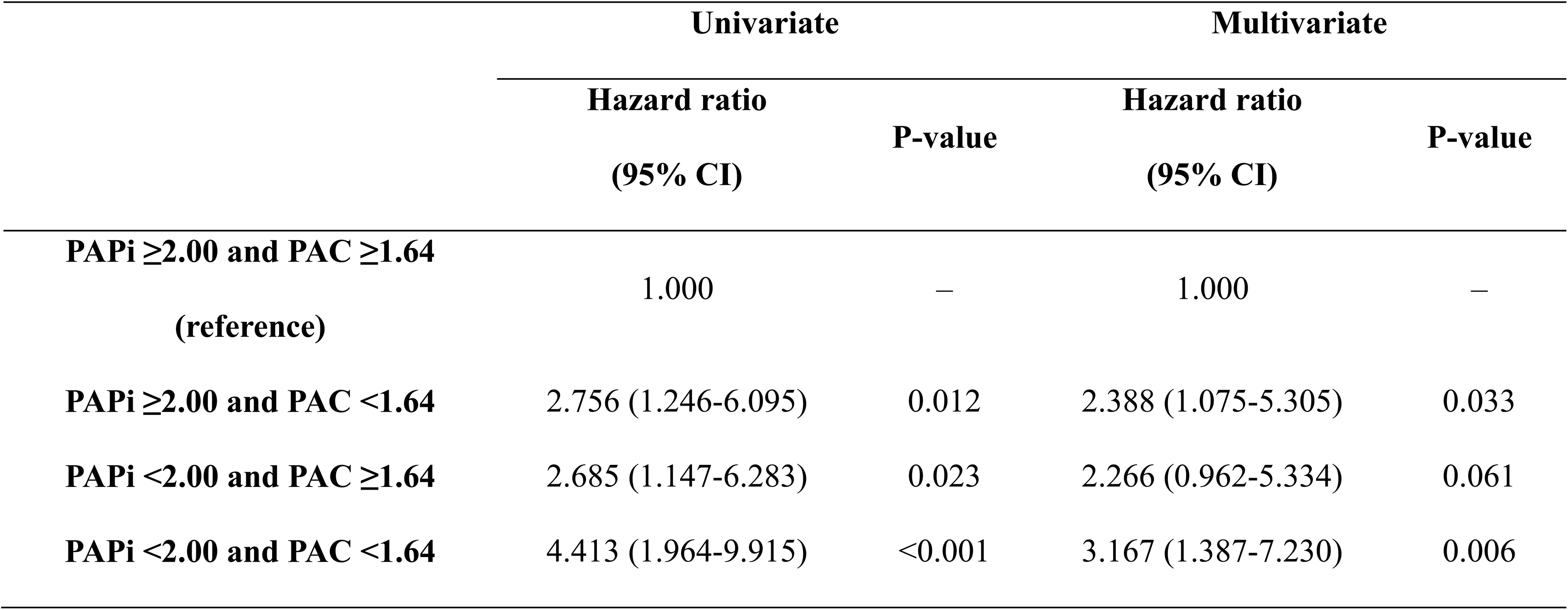
Association between hemodynamic phenotypes defined by the PAPi and PAC and the composite endpoint. Hazard ratios and 95% CIs were estimated using Cox proportional hazards models for the composite endpoint comprising all-cause death, left ventricular assist device implantation, or heart transplantation within 6 months. Patients were stratified into four groups based on bootstrap-derived optimal cutoffs for the PAPi (2.00) and PAC (1.64). Multivariable models were adjusted using the Get With The Guidelines-Heart Failure (GWTG-HF) risk score. CI, confidence interval; PAC, pulmonary arterial capacitance; PAPi, pulmonary artery pulsatility index.

**Table 3.**
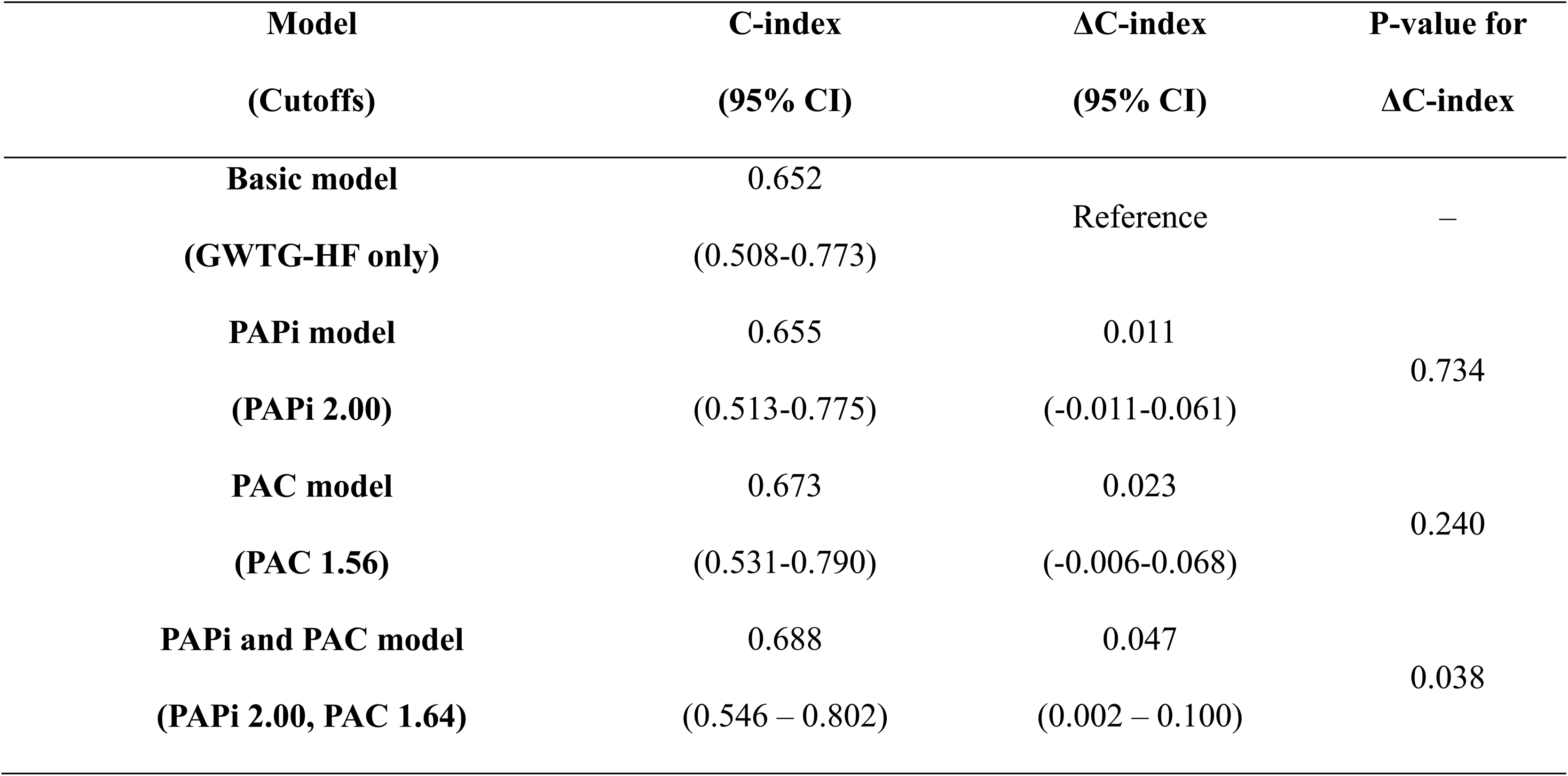
Incremental prognostic value of hemodynamic models based on PAPi and PAC. All models were adjusted for the GWTG-HF risk score. The composite endpoint was defined as all-cause death, left ventricular assist device implantation, or heart transplantation within 6 months. The C-index and 95% CIs were calculated to assess model discrimination. The basic model included only the GWTG-HF risk score. Incremental models incorporated dichotomized values of the PAPi and/or PAC. Cutoff values for PAPi (2.00) and PAC (1.64 for the combined model, 1.56 for the PAC-only model) were derived using a grid search procedure with bootstrap iterations, selecting values most frequently associated with the highest C-index. CI, confidence interval; GWTG-HF, Get With The Guidelines-Heart Failure; PAC, pulmonary arterial capacitance; PAPi, pulmonary artery pulsatility index.

Kaplan–Meier analysis demonstrated clear separation of clinical outcomes among the four hemodynamic groups (Figure 2). The low PAPi/low PAC group showed the lowest event-free survival, while the high PAPi/high PAC group exhibited the highest survival (log-rank P = 0.002).

**Figure 2.**
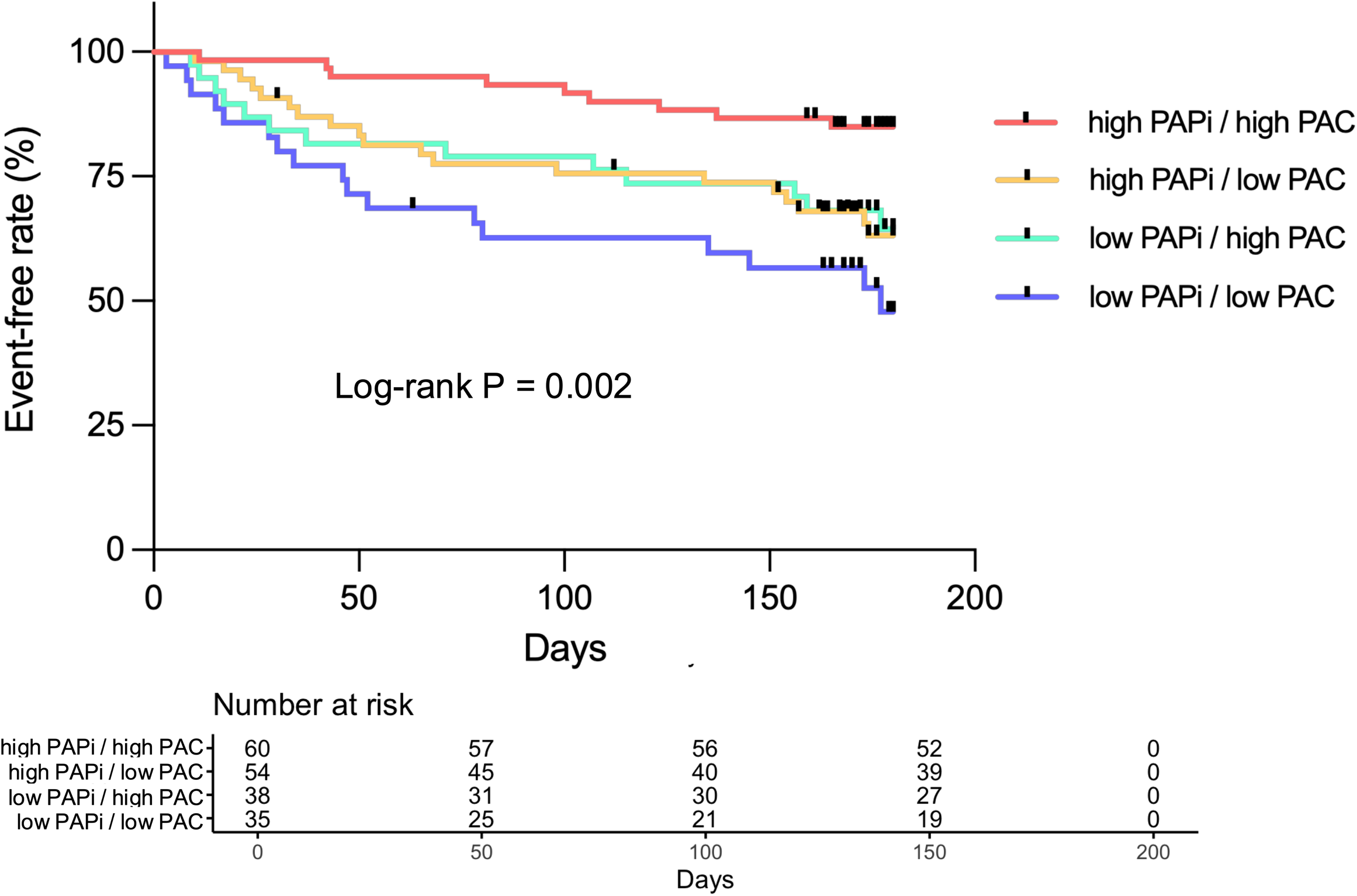
Kaplan–Meier curves stratified by hemodynamic groups defined by PAPi and PAC. Kaplan–Meier curves illustrating the event-free survival from the composite endpoint comprising all-cause death, left ventricular assist device implantation, or heart transplantation within 6 months, stratified by four hemodynamic groups defined by the PAPi and PAC. Patients were classified using bootstrap-derived optimal cutoffs (PAPi = 2.00 and PAC = 1.64) PAC, pulmonary arterial capacitance; PAPi, pulmonary artery pulsatility index.

In the multivariable Cox regression analysis adjusted for the GWTG-HF risk score, the low PAPi/low PAC group exhibited a significantly increased risk of the composite outcome (HR, 3.167; 95% CI, 1.387–7.230; P = 0.006) compared with the high PAPi/high PAC group; additionally, a high PAPi/low PAC was associated with a higher risk (HR, 2.388; 95% CI, 1.075–5.305; P = 0.033). Although the low PAPi / high PAC group demonstrated an elevated risk, this did not reach statistical significance (HR, 2.266; 95% CI, 0.962–5.334; P = 0.061).

In the analysis to evaluate the incremental prognostic value of the PAPi-PAC classification, the baseline model incorporating only the GWTG-HF risk score yielded a C-index of 0.652. The addition of the PAPi-PAC classification—constructed using grid search-derived cutoffs (PAPi 2.00, PAC 1.64)—improved model discrimination, increasing the C-index to 0.688 (ΔC-index, +0.047; 95% CI, 0.002–0.100; P = 0.038). Similarly, optimal cutoffs for PAPi and PAC were independently determined. The addition of the PAPi-based classification (cutoff 2.00) increased the C-index to 0.655 (ΔC-index, +0.011; 95% CI, −0.011–0.061; P = 0.734), while the addition of the PAC-based classification (cutoff 1.56) increased it to 0.673 (ΔC-index, +0.023; 95% CI, −0.006–0.068; P = 0.240). These results indicate that only the combined assessment of PAPi and PAC provides a statistically significant improvement in prognostic discrimination.

## Discussion

This study, utilizing baseline data from the ESCAPE trial, demonstrates that the combined assessment of PAPi and PAC at the time of hospitalization allows for significant prognostic stratification in patients with decompensated advanced HF. Importantly, this hemodynamic classification—based on early invasive measurements—was demonstrated to improve risk prediction beyond that achieved by a validated clinical risk score (15, 16).

The clinical utility of combining PAPi and PAC lies in their complementary role in reflecting right heart pathophysiology. RV function is highly sensitive to afterload, making its interpretation challenging in isolation (5–7). Accordingly, simultaneous assessment of RV function and afterload provides a more physiologically coherent and prognostically informative framework. In the present study, this combined approach offered superior risk discrimination compared with either metric alone, underscoring the incremental prognostic value of jointly evaluating RV function and afterload. These findings are consistent with our prior investigations, which independently demonstrated the prognostic value of the PAPi-PAC classification in HF with preserved and reduced ejection fraction, and extend the utility of this approach to include decompensated advanced HF (12, 13). While a prior sub-analysis of the ESCAPE trial identified baseline PAPi as an independent predictor of adverse outcomes, it did not assess model discrimination (9). Our results suggest that although PAPi may have individual prognostic value, its clinical interpretation is substantially enhanced when integrated with PAC, emphasizing the value of a composite RV-pulmonary vascular assessment.

The present findings suggest that early hemodynamic phenotyping using PAPi and PAC at the time of hospitalization allows for valuable prognostic stratification in patients with decompensated advanced HF—a clinical context in which invasive monitoring is commonly employed. This classification may help guide the timing and intensity of advanced therapies, including decision-making regarding durable mechanical circulatory support implementation or transplantation. In particular, patients with both low PAPi and low PAC exhibited the worst outcomes. These individuals exhibited elevated filling pressures and a reduced cardiac output, suggestive of advanced biventricular dysfunction with progressive pulmonary vascular remodeling. Such patients may represent a uniquely high-risk subgroup within the advanced HF population who could benefit from more aggressive or earlier intervention. Conversely, more conservative or stepwise management may be feasible in those with preserved PAPi and/or PAC. Accordingly, this stratification framework may aid clinicians in tailoring acute treatment strategies based on individualized hemodynamic risk.

Some limitations of this study should be noted. First, although the data were derived from a peer-reviewed randomized controlled trial (ESCAPE), the present analysis was retrospective in nature, prohibiting definite causal inferences. Second, the ESCAPE trial was conducted between 2000 and 2003, prior to the widespread adoption of contemporary HF therapies such as angiotensin receptor-neprilysin inhibitors and sodium-glucose cotransporter 2 inhibitors. Third, the PAPi and PAC thresholds used for risk stratification were derived using a bootstrap-based grid search algorithm within this dataset. While this approach enhanced internal consistency, external validation was not performed. Finally, the ESCAPE cohort exclusively included patients with advanced HF, and the generalizability of these cutoffs to a broader population with acute decompensated HF remains unclear.

In patients hospitalized for decompensated advanced HF, early hemodynamic assessment using the PAPi and PAC provided incremental prognostic value beyond established clinical risk scores. This stratification approach identified distinct phenotypes associated with differential risks of death, durable mechanical circulatory support implementation, or heart transplantation. These findings may help guide timely considerations regarding advanced therapies in patients at a higher risk.

## Data Availability

The data that support the findings of this study are available from the National Heart, Lung, and Blood Institute's Biologic Specimen and Data Repository Information Coordinating Center but restrictions apply to the availability of these data, which were used under license for the current study, and so are not publicly available. Data are, however, available from the authors upon reasonable request and with permission from the National Heart, Lung, and Blood Institute's Biologic Specimen and Data Repository Information Coordinating Center.

## Acknowledgments

The authors acknowledge the National Heart, Lung, and Blood Institute and the Biologic Specimen and Data Repository Information Coordinating Center for providing access to the Evaluation Study of Congestive Heart Failure and Pulmonary Artery Catheterization Effectiveness trial dataset. The data utilized in this study were obtained from the Blood Institute and the Biologic Specimen and Data Repository Information Coordinating Center for the purposes of the current analysis, based on an approved data request. The authors are solely responsible for the design, analysis, interpretation, and conclusions of this study, which do not necessarily represent the views of the National Heart, Lung, and Blood Institute or the Blood Institute and the Biologic Specimen and Data Repository Information Coordinating Center.

## Sources of Funding

None.

## Disclosures

None.

## Non-standard Abbreviations and Acronyms

C-index: concordance index
CI: confidence interval
ESCAPE: Evaluation Study of Congestive Heart Failure and Pulmonary Artery Catheterization Effectiveness
GWTG-HF: Get With The Guidelines-Heart Failure
HF: heart failure
HR: hazard ratio
PAC: pulmonary arterial capacitance
PAPi: pulmonary artery pulsatility index
RV: right ventricular

## Notes

### Competing Interest Statement

The authors have declared no competing interest.

### Author Declarations

Ethical approval was waived by the institutional review board of the Anjo Kosei Hospital ethic committee on April 22, 2025 (Waiver No. R24-031), due to the retrospective nature of the study using de-identified data.

